# Chronic thromboembolic pulmonary hypertension in UAE: the first reported seven year outcomes from a tertiary center in UAE

**DOI:** 10.1101/2023.11.05.23298119

**Authors:** Khaled Saleh, Jihad Mallat, Kelly Dougherty, Mohannad Ghanem, Ahmed Ghorab, Reem Alsaadi, Manyoo Agrawal, Simi Salim, Sara Abdalla, Naureen Khan, Hani Sabbour

**Affiliations:** Respiratory Institute, Cleveland Clinic Abu Dhabi, Abu Dhabi P.O. Box 112412, United Arab Emirates; Critical Care Institute, Cleveland Clinic Abu Dhabi, Abu Dhabi P.O. Box 112412, United Arab Emirates; Cleveland Clinic Lerner College of Medicine, Case Western Reserve University, Cleveland, OH 44106, USA; Normandy University, UNICAEN, ED 497, 14032 Caen, France; New York University Langone Health, Grossman Medical School, NY 11238; School of Medicine Royal College of Surgeons in Ireland, Bahrain; Medical subspeciality Institute, Cleveland Clinic Abu Dhabi, Abu Dhabi P.O. Box 112412, United Arab Emirates; Heart and Vascular Institute, Cleveland Clinic Abu Dhabi, Abu Dhabi P.O. Box 112412, United Arab Emirates; Jawaharlal Institute of Postgraduate Medical Education and Research, Puducherry, India; Cardiology SPR, Basingstoke Hospital NHS, UK; St. George’s University, West Indies, Grenada

**Keywords:** chronic thromboembolic pulmonary hypertension, six-minute walk test, N-terminal pro-brain natriuretic peptide, pulmonary arterial wedge pressure, pulmonary vascular resistance, right systolic ventricle pressure, cardiac index, world health organization

## Abstract

**Background:** The aim of the study was to present the first United Arab Emirates (UAE) Chronic Thromboembolic Pulmonary Hypertension registry of patients’ clinical characteristics, hemodynamic parameters, and treatment outcomes.

**Methods:** This was a retrospective study describing all adult patients who were diagnosed with Chronic Thromboembolic Pulmonary Hypertension (CTEPH) between January 2015 and April 2022 in a tertiary referral center in Abu Dhabi, United Arab Emirates (UAE).

**Ethics Statement:** IRB and Research Ethics Committee approval was obtained under REC number A-2017-030. Researchers had access to the anonymized electronic medical records with fully anonymized data for analysis of retrospective data collected from July 2015 till April 2022. IRB and REC waived the requirement for informed consent.

**Results:** A total of 39 consecutive patients were diagnosed with CTEPH during the seven years of the study. Two patients who had pulmonary artery balloon angioplasty (BPA) were not included in the analysis. Twelve out of the 37 patients (56%) had pulmonary thrombo-endarterectomy (PTE) and were designated as the surgical group, the rest were designated the medical group. The patients were followed for a median of 11 months [IQR: 6-22.5]. Most of the patients in the medical group were started on dual then subsequently escalated to triple combination therapy. Most of the patients in the surgical group stopped the vasodilator therapy. The 1-, 3- and 5-year medical group survival rates were 80%, 68% and 68%, respectively. None of the patients in the surgical group died.

**Conclusion:** This is the first registry of CTEPH patients from a single tertiary referral center in UAE. Our cohort was younger than ones reported in the Western World but similar to the one reported in Saudi Arabia. The medical group mortality is comparable to other major registries. The recent introduction of PTE program in our center is likely to increase number of surgical patients.

## Introduction

Chronic thromboembolic pulmonary hypertension (CTEPH) is a condition where pulmonary hypertension (PH) occurs due to pulmonary artery obstruction from unresolved thromboembolic disease which can lead to right ventricular failure and ultimately death ^1^. Our understanding of CTEPH characteristics and clinical outcome data have been published in Europe ^2^, North American ^3^, Middle East ^4^, and Far East ^5^. Recent advancement has been done in the diagnosis and management of CTEPH from newly approved medication to treat the pulmonary hypertension to relief of the obstruction mechanically by pulmonary balloon angioplasty or surgical removal by thromboendarterectomy ^1^. There is a paucity of data on CTEPH clinical characteristics and outcomes of patients in the Middle East and North African region. We present the first data from United Arab Emirates, defining baseline characteristics and comparison of medical treatment vs. surgical pulmonary thrombo-endarterectomy (PTE) outcome. The surgical intervention was mostly done in Europe and the United States of America (USA) due to lack of availability during this study period.

## Method

This was a single center retrospective study of consecutive patients over the age of 14 years at Cleveland Clinic Abu Dhabi who were known or suspected of having PH between January 2015 and April 2022. Exclusion criteria included patients who had pulmonary arterial hypertension causes other than CTEPH, acute pulmonary embolism or if patient did not have right heart catheterization (RHC). All patients had an echocardiography, N-terminal pro B-type natriuretic peptide (NT-proBNP) and 6 minutes-walk test (MWT). All patients had ventilation perfusion scan in addition to CT angiogram of the chest to confirm the disease. All patients had complete pulmonary function tests, abdominal ultrasound, hepatic virology screen, liver function tests, and serologies for connective tissue diseases or hyper-coagulable states when suspected. A diagnostic RHC was required for the diagnosis of PH and to be included in the study. If a wedge could not be measured, the left ventricular end-diastolic pressure was directly measured.

The study protocol was approved by the Research Ethics Committee at Cleveland Clinic Abu Dhabi (REC Number: A-2017-030). The clinical characteristics included age, sex, and body mass index (BMI). The classification of PAH was based on definitions outlined in the previous 2018 6^th^ World Symposium of Pulmonary Arterial Hypertension(Galiè et al., 2019). PAH was defined by mean pulmonary artery pressure (mPAP) ≥ 25 mmHg, wedge pressure (WP) ≤ 15 mmHg and pulmonary vascular resistance (PVR) ≥ 3 woods unit. Clinical data for PAH risk stratification was collected as recommended by the 6^th^ World symposium including World Health Organization functional class (WHO FC), NT-proBNP, 6-MWT in meters.

Pulmonary vasodilator medications were reported on the first and last visits. The medications used were phosphodiesterase-5 inhibitors (sildenafil or tadalafil) or soluble guanylate cyclase inducer (riociguat) usually started as monotherapy, and endothelin receptor antagonists (ambrisentan or macitentan) usually added as second medication. Prostacyclin analogs selexipag, nebulized iloprost or subcutaneous/intravenous prostacyclin treprostenil were used as third medication when clinically indicated. We used the Simplified French model as a non-invasive risk stratification tool for follow up visits, utilizing 3 low-risk criteria: WHO FC I/II, 6 MWT >440 meters, and NT-proBNP <300 ng/L ^8, 9^.

### Statistical analysis

All data are expressed as median [25-75%, interquartile range, [IQR] since the sample size is small. Comparisons of continuous variables between different groups or subgroups were performed by Wilcoxon rank sum test. Pairwise comparisons between different study times for continuous variables were assessed using Wilcoxon signed rank test. Comparisons of categorical data between the different groups were performed using the Chi2 or Fisher’s exact tests, as appropriate. Pairwise comparisons between different study times for categorical variables were assessed using McNemar’s test. The cumulative probability of survival was analyzed using the Kaplan-Meier method. Patients who did not experience the event (death) during the study period were censored at the time of last seen. Differences between survival curves were assessed using the log-rank test.

Statistical analysis was performed using STATA 14.0 (StataCorp LP, College Station, Texas, USA). P <0.05 was considered statistically significant. All reported P values are 2-sided.

## Results

During the study period, a total of 39 patients were diagnosed with CTEPH. Two patients who had pulmonary artery balloon angioplasty (BPA), one in Vienna and another in Cleveland Clinic Ohio, USA, were excluded from the study due to small number. They both had resolution of the pulmonary hypertension and were weaned off vasodilator therapy. The clinical characteristics and hemodynamic findings of the 37 patients are shown in Table 1. The patients were followed for a median of 11.0 months [IQR: 6.0-22.5]. The majority were Emirati nationals (62%) with the remaining from the middle east and south Asia. The majority were females (68%). Three patients had hyper-coagulopathy (lupus anticoagulant) as the most common encountered risk factor while one patient had factor V Leiden deficiency. About half of the patients (46%) were obese with a median BMI of 30.0 [IQR: 24.0-35.4].

**Table 1.** Baseline demographic, clinical characteristics and hemodynamics.

| Variable | All patients<br>(n=37) | Medical<br>(n=25) | Surgical<br>(n=12) | P-Value |
| --- | --- | --- | --- | --- |
| Age, yr | 46 [33-59] | 46 [34-61] | 44 [32-53] | 0.592 |
| Female, n (%) | 25 (68) | 17 (68) | 8 (67) | 0.935 |
| Emirati, n (%) | 23 (62) | 14 (56) | 9 (75) | 0.256 |
| BMI, kg/m<sup>2</sup> | 30.0 [24.0-35.4] | 28.1 [22.9-34] | 33.5 [27.2-40.9] | 0.102 |
| Follow-up, month | 11.0 [6.0-22.5] | 11.0 [5.0-19.0] | 13.5 [8.5-27.0] | 0.36 |
| Baseline 6-MWT, m | 311 [237-362] | 284 [237-350] | 331 [189-426] | 0.640 |
| Baseline NT-proBNP, pg/mL | 600 [139-1467] | 912 [139-2388] | 292 [126-902] | 0.090 |
| WHO III/IV, n (%) | 29 (78.4) | 19 (76) | 10 (83) | 0.606 |
| Mortality % | 6 (16) | 6 (24) | 0 (0) | 0.149 |
| mRAP, mmHg | 7 [4-10] | 7 [4-11] | 9 [6-10] | 0.675 |
| mPAP, mmHg | 46 [27-52] | 45 [31-52] | 48 [22-52] | 0.741 |
| SPAP, mmHg | 79 [41-87] | 76 [50-88] | 79 [36-85] | 0.555 |
| DPAP, mmHg | 21 [13-32] | 22 [14-34] | 18 [10-33] | 0.524 |
| PAWP, mmHg | 10 [7-12] | 9.5 [7-11] | 12 [7-21] | 0.219 |
| CI (Fick method), L/min/m<sup>2</sup> | 2.1 [1.9-2.5] | 2.1 [1.9-2.6] | 2.1 [2-2.5] | 0.739 |
| PVR (Fick method), mmHg/L/min | 621 [223-995] | 651 [260-1086] | 588 [199-769] | 0.505 |
| SVO<sub>2</sub>, % | 60 [52-70] | 57 [51-72] | 68 [56-68] | 0.650 |
| RSVP, mmHg | 61 [50-75] | 66 [58-87] | 54[25-72] | 0.089 |

BMI, body mass index; 6-MWT, six-minute walk test; NT-proBNP, N-terminal pro-brain natriuretic peptide; TAPSE, tricuspid annular plane systolic excursion; WHO, world health organization; mRAP, mean right atrial pressure; mPAP, mean pulmonary arterial pressure; PAWP, pulmonary arterial wedge pressure; CI, cardiac index; PVR, pulmonary vascular resistance; SVO<sub>2</sub>, venous oxygen saturation; LVEF, left ventricle ejection fraction; RSVP, right systolic ventricle pressure.
Data are expressed as median and interquartile [25-75] or count and percentage (%).

Twelve out of the 37 patients (56%) had PTE and were designated as the surgical group, the rest were designated the medical group. The clinical and hemodynamic characteristics were not significantly different between the two groups (Table 1). Over the follow up period, the surgical group had significant improvement in NT-proBNP and WHO FC compared to the medical group. There was a trend but not statistically significant improvement in 6-MWT and right ventricular systolic pressure (Table 2).

**Table 2.** Changes in clinical, echocardiographic parameters and low clinical risk criteria at diagnosis and last follow-up visit.

| Variables | Medical (n=25) |  |  | Surgical (n=12) |  |  |
| --- | --- | --- | --- | --- | --- | --- |
|  | Baseline | Last visit | P-value | Baseline | Last visit | P-value |
| 6-MWT, m | 284 [237-349] | 315 [241-448] | 0.18 | 331[188-425] | 350 [329-469] | 0.09 |
| NT-proBNP, pg/mL | 912 [139-2388] | 3178 1062[95-3178] | 0.30 | 292[126-902] | 194 [94-232] | **0.04** |
| WHO III/IV, n (%) | 19 (76) | 12 (48) | 0.12 | 10 (83) | 2 (16.7) | 0.008 |
| RSVP, mmHg | 66 [58.5-87] | 73 [40-81] | 0.25 | 54 [25-72] | 31 [20-42] | 0.09 |
| Low clinical risk criteria |  |  |  |  |  |  |
| None, n (%) | 15 (60.0) | 13 (52.0) | 0.62 | 4 (33.3) | 1 (8.3) | 0.25 |
| One, n (%) | 7 (28.0) | 6 (24) | 1.00 | 3 (25.0) | 2 (16.7) | 1.00 |
| Two, n (%) | 0 (0) | 1 (4.0) | NA | 4 (33.3) | 5 (41.7) | 1.00 |
| Three, n (%) | 2 (8.0) | 2 (12.0) | 1.00 | 0 (0.0) | 4 (33.3) | 0.12 |

6-MWT, six-minute walk test; NT-proBNP, N-terminal pro-brain natriuretic peptide; WHO, world health organization; NA, not applicable.
Data are expressed as median and interquartile [25-75] or count and percentage (%).

Ten of the patients (40%) in the medical group and 3 (25%) in the surgical group had repeat RHC, which was done before the escalation of therapy following clinical deterioration.

On the last reported visit, the medical group had 13 (52.0%), 6 (24.0%), 1 (4.0%), and 2 (12.0%) patients in the zero, one, two and three low clinical risk criteria while the surgical group had 1 (8.3%), 2 (16.7%), 5 (41.7%), and 4 (33.3%) patients, respectively, based on the Simplified French model (Table 2).

During the follow up period, six patients (24%) died in the medical group. Two patients died within six months of diagnosis as they presented with advanced disease and were not surgical candidate and a third patient had massive pulmonary embolism as she was not adherent to anticoagulation and presented in cardiogenic shock with failed thrombolytic therapy. The remaining three died as complication of CTEPH. The 1-, 3- and 5-year survival rates were 86.4% (95%CI:67.8-94.7), 68.0 (95%CI:35.0-86.7) and 68.0% (95%CI:35.0-86.7), respectively. None of the patients in the surgical group died (Figure 2).

## Discussion

Our study is the first report of CTEPH patients’ outcome in the UAE. Patients were diagnosed and treated based on published guidelines ^10^. Of the twelve patients who had PTE, 7 were performed in Vienna, 3 in the UK and one in each of the USA and France since neither surgical PTE nor balloon pulmonary angioplasty were available at any center in the UAE.

Six of the 25 medical patients were surgical candidates but did not have financial support to travel to perform the surgery. All the patients in the medical group had escalation of vasodilator therapy with the majority requiring triple therapy (Figure 1). On the first visit, most of the patients on monotherapy were on riociguat considering it is the only approved medication for CTEPH. Escalation to dual therapy was done by adding endothelin receptor antagonists-(ERA) macitentan or ambrisentan. The triple therapy included subcutaneous treprostinil or oral prostacyclin analog, selexipeg. Some patients in the clinical high-risk ERS criteria were treated with selexipeg since intravenous or subcutaneous form was not covered by their insurances.

**Figure 1.**
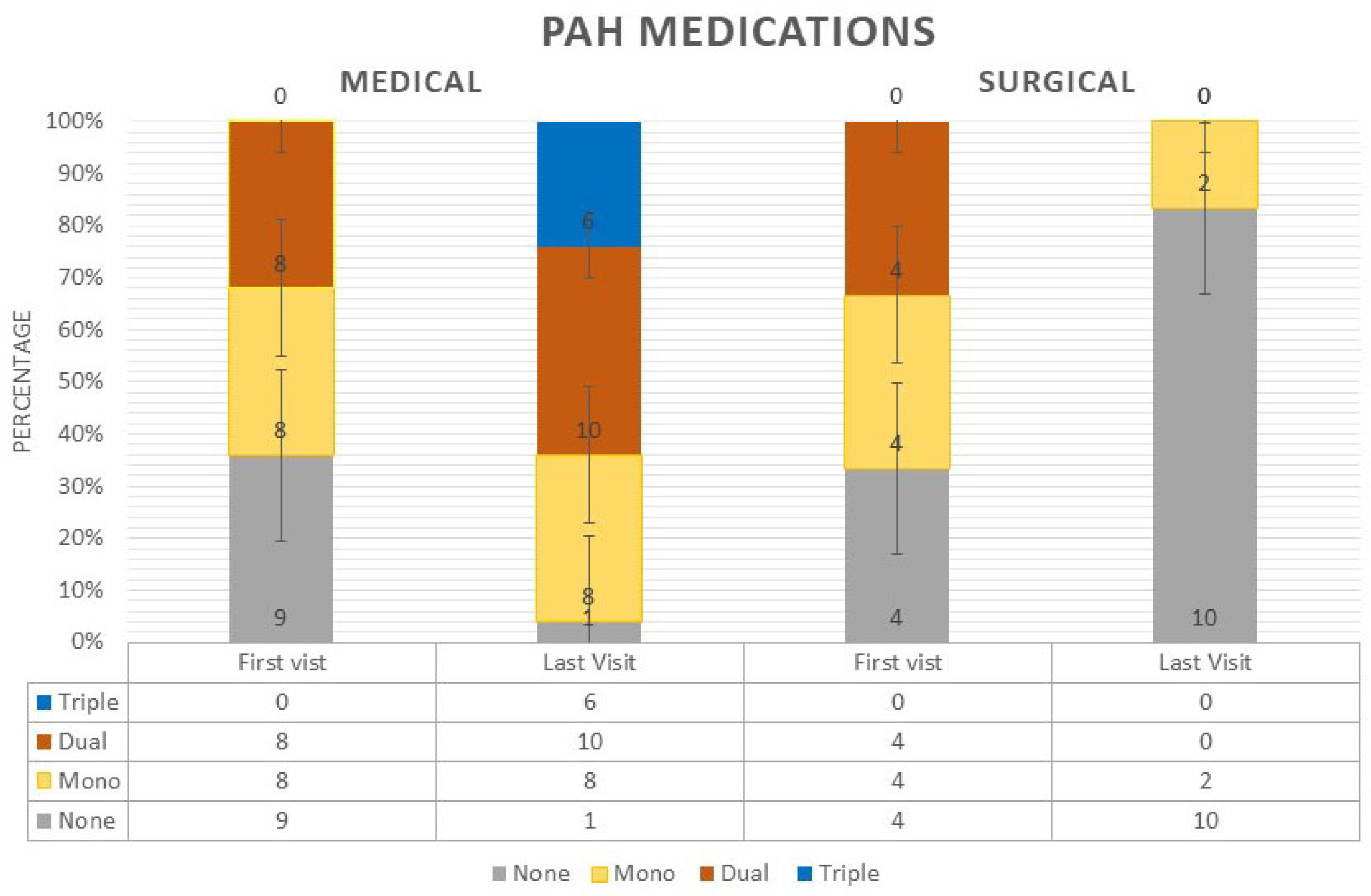
None, mono, dual and triple vasodilator therapies for Medical and Surgical group first and last reported visits.

**Figure 2.**
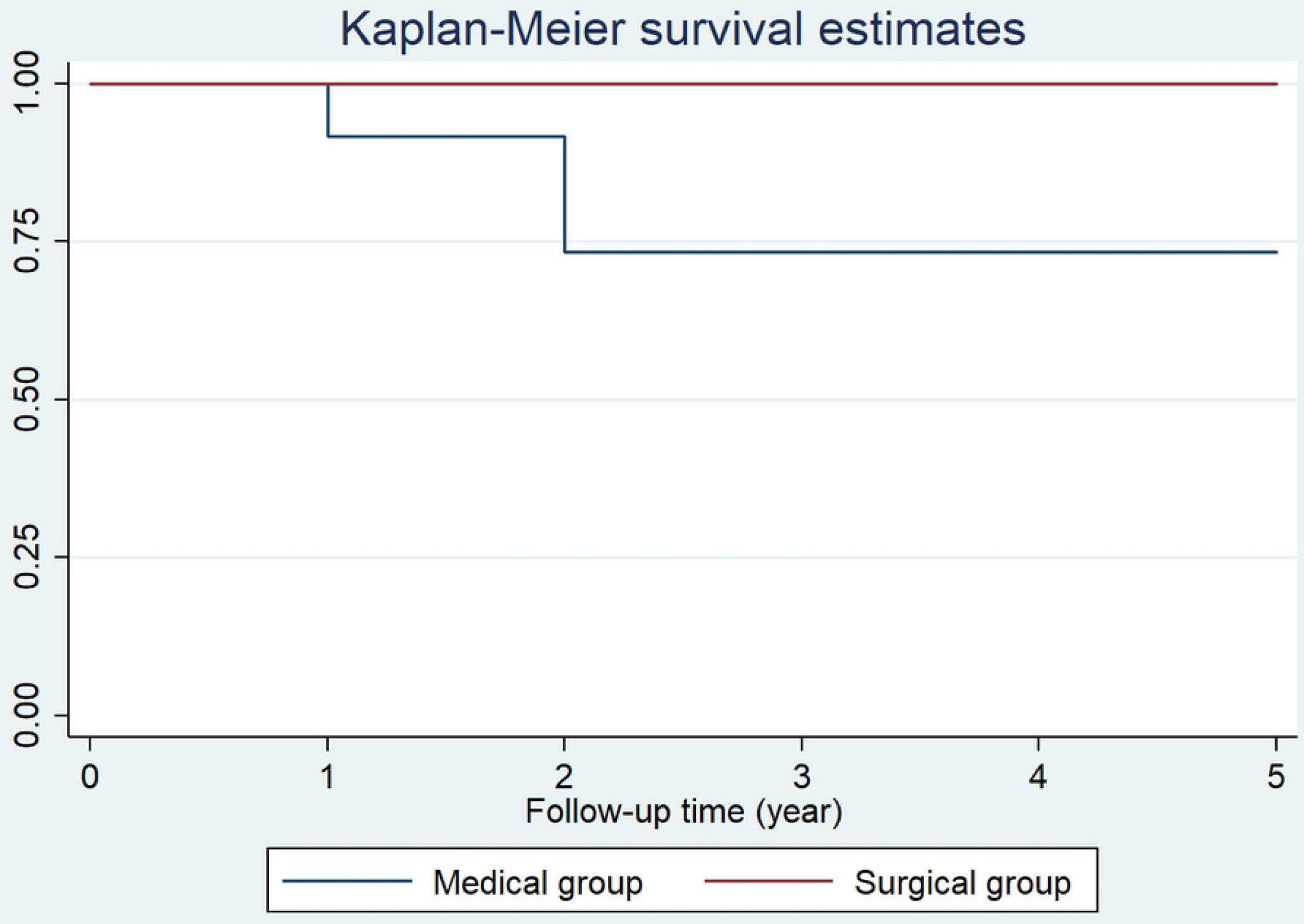
Survival for the Medical and Surgical group over the study period.

Four of the 12 patients (33.3%) in the surgical group did not receive bridging vasodilator therapy. Four were on monotherapy and 4 were on dual therapy prior to the surgery. Post operatively, ten of the twelve patients were able to stop the vasodilator therapy except 2 were maintained on riociguat. Most of the patients in the surgical group ended in the two or three low clinical risk criteria categories while most patients in the medical group ended in the zero low clinical risk criteria indicating a worse prognosis for the medical group (Table 2). This is in relative agreement with the International Registry in terms of significant clinical improvement in patients who had PTE ^2,11^.

Our 1 and 3-years survival for the medical group were 86.4% and 68.0%, respectively, which is not significantly different from the 92% and 75% of the Compera Registry ^11^, 89% and 74% of the subgroup in the French registry ^12^ as well as the older German and UK registries ^13, 14^. One study from Saudi Arabia registry reported only 1-year survival in the medical group of 94.4% ^4^. Despite having significantly smaller number of patients compared to the above registries, six of our patients (25%) had triple therapy including prostacyclin analogue compared to 2 % in the French registry. While we had four patients on selexipag as third added oral medication, there was only one patient in the Saudi registry. Our patient population were younger than the European registries (mean age 68) but similar to the Saudi Registry mean age of 40 (SD 11). Most of our patients were also in WHO FC class III and IV (75%) similar to the above-mentioned registries ^2, 4, 12, 13^.

The surgical group had 100 % 1 and 3-year survival rates. There were no post operative complications and none of the patients had any significant residual pulmonary hypertension based on the clinical findings. We stopped vasodilator medications on all except two patients who remained stable (low risk Criteria) on riociguat, prescribed based on current evidence ^15^. Considering the small number of patients who had PTE done in different centers of excellence in Europe and the USA, no comparison was done with the results of other registries. The choice of the centers was based on the cost and surgical outcome decided solely by the payor. We have since started a new surgical PTE program at our hospital, the outcome of which will be reported in the future.

The limitations of the study include the retrospective nature and the small number of patients. Many patients who could be surgical candidate could not have the surgery due to the lack of insurance coverage. Also, the PTE surgeries were done at different major centers of excellence in Europe and the USA, which could have biased the surgical outcomes.

## Conclusion

This was the first registry of CTEPH patients from a single tertiary referral center in UAE. Our patients were younger than the reported registries in Europe and the USA but similar to the one reported in Saudi Arabia. Our mortality outcome in the medically treated group is comparable to other major registries around the world. The excellent surgical outcome reflects the detailed preoperative evaluation, management and appropriate patients’ selection. It additionally reflects the expected procedural outcomes of major CTEPH centers of excellence in Europe and the USA.

## Data Availability

All the relevant data are within the manuscript and its supporting information files

NA

## Abbreviations

CTEPH: chronic thromboembolic pulmonary hypertension
BPA: pulmonary artery balloon angioplasty
UAE: United Arab Emirates
PTE: pulmonary thrombo-endarterectomy
PH: pulmonary hypertension
USA: United States of America
NT-proBNP: N-terminal pro B-type naturitic peptide
6-MWT: 6 minutes-walk’s test
RHC: right heart catheterization
BMI: body mass index
mPAP: mean pulmonary artery pressure
WP: wedge pressure
PVR: pulmonary vascular resistance
WHO FC: World Health Organization functional class

## Contributions

Study concept: KS and HS

Data collection: KD, RA, SA, SS, and AG

Data analysis: KS, HS, and JM

Statistical analysis: JM

Discussion of results: KS, HS, and JM

Drafting of the manuscript: KS, HS, and JM

Revision of the final manuscript: KS, MG, AG, MA, JM, NK, and HS

All authors read and approved the manuscript.

## Ethics approval

This study was approved by the Cleveland Clinic Abu Dhabi Ethics Committee (REC Number: A-2017-030).

